# Scalp Bacterial Microbiota Dysbiosis in Androgenetic Alopecia: Community Structure, Functional Profiles, and Associations with Lifestyle Factors

**DOI:** 10.64898/2026.06.24.26356478

**Authors:** Yiwen He, Lulu Zhu, Dengfeng Lv, Juntong Yu, Jiafeng Yang, Jinyu Wu, Jiale Jin, Gang Deng

## Abstract

The aim of this study was to explore the scalp bacterial flora structure and functional characteristics in androgenetic alopecia (AGA) patients, analyze its association with disease phenotypes and unhealthy lifestyles, and provide a basis for clarifying AGA’s microecological pathogenic mechanism and targeted interventions. A total of 7 AGA patients and 6 healthy controls (HC) were enrolled, with scalp microbial samples collected. High-throughput sequencing of the 16S rRNA V3–V4 region was used to analyze flora alpha/beta diversity, species composition and differential species. LEfSe and KEGG functional prediction screened marker bacteria and differential pathways, and clinical/lifestyle data were collected for inter-group comparisons. No significant difference in Chao index was observed between groups (*P*>0.05), but Shannon/Simpson indices/Pielou evenness (*P*<0.01) and intra-group Bray–Curtis distance (*P*<0.001) were significantly higher in the AGA group, indicating reduced community stability. *Staphylococcus* dominated healthy scalps; the AGA group had fewer symbiotic bacteria but enriched *Acinetobacter*, *Pseudomonas*, and *Cutibacterium*. LEfSe identified *Firmicutes*/ *Staphylococcus* as HC markers and *Proteobacteria*/*Gammaproteobacteria*/ *Acinetobacter*/*Pseudomonas* as AGA dysbiotic flora. KEGG showed upregulated metabolic, immune and cell motility pathways in AGA (*P*<0.05), with only infectious diseases pathway enriched in HC. AGA patients had more frequent hair washing and higher rates of staying up late, high-fat diet and insufficient fruits/vegetables (all *P*<0.05). In conclusion, AGA patients have typical scalp microecological dysbiosis closely related to unhealthy lifestyles, which may accelerate alopecia by inducing follicular inflammation. Scalp flora can be potential biomarkers and targets for AGA assessment and intervention.

## Introduction

Androgenetic alopecia (AGA) is a non-scarring, progressive alopecic disease characterized by hair follicle miniaturization, which mostly starts in adolescence and is the most common type of alopecia in clinical practice. According to statistics, approximately 65% of the global population is affected by it, among which the prevalence rate is about 21.3% in Chinese men and 6.0% in Chinese women, while the incidence rate in white men is as high as 80%[1–3]。

The occurrence and development of AGA are synergistically regulated by multiple factors such as heredity, hormones and environment. Among them, genetic susceptibility and abnormal androgen metabolism are the core pathogenic factors, while the impact of environmental factors cannot be ignored[4, 5] 。 Contemporary people generally have problems such as irregular diet, staying up late and insomnia, and frequent mood swings, leading to an annual increase in the prevalence of AGA and a younger age of onset. This disease not only affects the external image of patients, but also easily causes negative emotions such as anxiety and inferiority, seriously reducing their quality of life[6, 7]。

Human skin and hair are important hosts for a variety of microorganisms. The balanced state of the scalp microecology is crucial for the health of the skin and hair, and its imbalance is closely related to the occurrence of various skin and hair diseases[8, 9]。Previous studies have confirmed that there is obvious disorder in the scalp microecology of AGA patients, and there are significant differences in the abundance of dominant flora and community diversity compared with healthy people[10, 11] 。 This microecological imbalance may further accelerate the process of hair follicle miniaturization by inducing local inflammatory responses of the scalp, interfering with normal hair follicle metabolism, and damaging the scalp barrier function, thereby participating in the occurrence and development of AGA[12]。

In this study, 7 AGA patients were selected as the alopecia group and 6 healthy people as the control group. By investigating the basic demographic information, dietary and living habits, and scalp status of the two groups of subjects, combined with 16S rRNA V3-V4 region sequencing technology, the differences in community composition and diversity of scalp microecology between the two groups were systematically analyzed, and the internal correlation between scalp microecology and AGA was explored. This study provides basic experimental basis for improving the theoretical mechanism of AGA pathogenesis and exploring individualized intervention schemes based on microecological regulation.

## Materials and Methods

### Subject Recruitment and Sample Collection

A total of 13 volunteers aged 18–60 years were recruited for this study between 15/05/2026 and 01/06/2026, among whom 6 healthy volunteers were assigned to the healthy control (HC) group and 7 volunteers with androgenetic alopecia (AGA) were assigned to the experimental group. The study was approved by the Ethics Committee of The First Affiliated Hospital of Ningbo University ((Ethics No. 2026-R189-01). All participants gave their written informed consent before taking part in the study. All methods were carried out in accordance with the approved protocol. Volunteers were screened in accordance with the inclusion and exclusion criteria and enrolled consecutively. The exclusion criteria were as follows: females who were planning pregnancy, pregnant, or lactating; individuals who had taken oral antibiotics, immunomodulators, special diets, or probiotic preparations within the past 1 month; individuals who had received topical or hormonal treatment on the scalp within the past 3 months; individuals who had undergone hair perming or dyeing within the past 3 months; and individuals with other skin diseases. A questionnaire survey was used to record the basic information of volunteers, including height, weight, and age, as well as to assess scalp conditions, living habits, and dietary habits.

All participants were required to avoid washing or otherwise treating the scalp within 48 hours prior to sampling. For sampling, the operators wore disposable sterile gloves, moistened sterile cotton swabs with sterile normal saline, and wiped the designated scalp regions horizontally and vertically for 30 seconds over an area of approximately 4 cm². The swab tips were then cut off and placed in sterile cryotubes, snap-frozen in liquid nitrogen, and stored at −80 °C until further analysis.

### Bacterial Genomic DNA Extraction and Quality Control

DNA was isolated using the HiPure DNA Kits (Magen Biotech, Guangzhou, China), following the manufacturer’s recommended protocol. The quality and quantity of the extracted genomic DNA were assessed using two complementary approaches. First, DNA concentration and purity were measured using a NanoDrop 2000 spectrophotometer (Thermo Fisher Scientific, Waltham, MA, USA) with 2 μL of the sample. The absorbance ratios A260/A280 and A260/A230 were determined to evaluate DNA purity; an A260/A280 ratio of 1.8–2.0 was considered indicative of pure DNA, while an A260/A230 ratio ≥ 1.8 was used as the threshold for acceptable levels of organic contaminants such as carbohydrates, phenols, or salts. Second, DNA integrity was verified by 1% (w/v) agarose gel electrophoresis. Intact genomic DNA was expected to appear as a distinct, single band with no significant smearing (indicating degradation) or additional bands (indicating protein or RNA contamination). Only DNA samples meeting all quality criteria were used for subsequent library preparation and sequencing.

### Illumina 16S Ribosomal RNA Gene V3–V4 Amplicon Sequencing Library Preparationand Sequencing

PCR amplification was performed with diluted genomic DNA as the template. Depending on the selected sequencing region, barcode-containing specific primers were employed to amplify the target regions of 16S rRNA, ITS, or 18S rRNA. For the V3-V4 hypervariable region of 16S rRNA, the primers used were 341F (sequence: CCTACGGGNGGCWGCAG) and 806R (sequence: GGACTACHVGGGTATCTAAT), with an expected amplicon length of approximately 466 bp. A 50 μL PCR amplification system was prepared with Q5 high-fidelity DNA polymerase, comprising 10 μL of 5×Q5 Reaction Buffer, 10 μL of 5×Q5 High GC Enhancer, 1.5 μL of 2.5 mM dNTPs, 1.5 μL of 10 μM forward primer, 1.5 μL of 10 μM reverse primer, 0.2 μL of Q5 High-Fidelity DNA Polymerase, X μL of template DNA (50 ng total), and nuclease-free sterile water to bring the final volume to 50 μL. PCR amplification was conducted on a quantitative thermal cycler under the following parameters: initial denaturation at 95 ℃ for 5 min; followed by 30 cycles of 95 ℃ denaturation for 1 min, 60 ℃ annealing for 1 min, and 72 ℃ extension for 1 min; and a final extension at 72 ℃ for 7 min. Post-amplification, PCR products were purified using AMPure XP Beads (Beckman, CA, USA), and the concentration of purified products was accurately quantified using a Qubit 3.0 Fluorometer. Sequencing libraries were subsequently constructed using the Illumina DNA Prep Kit (Illumina, CA, USA). Library quality was assessed using the ABI StepOnePlus Real-Time PCR System (Life Technologies, Foster City, USA), followed by library pooling and sequencing on the Illumina NovaSeq 6000 platform in PE250 mode.

### 16S rRNA Gene V3-V4 ASV Data Processing

Raw sequencing data were filtered using FASTP (v0.18.0) [13] to remove reads containing ≥10% unknown nucleotides (N), ≥50% bases with a Phred quality score ≤20, or adapter sequences, thereby obtaining clean reads. FLASH (v1.2.11)[14] was used to assemble clean reads into tag sequences (minimum overlap length 12 bp, maximum mismatch rate 2%); low-quality tags were filtered according to the criteria described in the literature [15, 16] and further processed following the Qiime analysis workflow (truncation of consecutive low-quality bases and removal of insufficient high-quality tags) to obtain high-quality clean tags. The DADA2 plugin (v1.14)[17] of Qiime2 was employed to remove primer sequences, construct a denoising error model, and generate raw amplicon sequence variants (ASVs); meanwhile, the UCHIME algorithm [18] was used to remove chimeric sequences, and finally, denoised and chimeric-free ASVs and their abundance information were obtained. Operational Taxonomic Unit (OTU) characteristics were summarized based on abundance and annotation data; Core and Pan OTU rarefaction curves were plotted using the OTU abundance table. OTU/ASV sequences were aligned against the SILVA (v138.1)[19] database, and species annotation was completed using the naive Bayesian model of the RDP software (v2.14)[20] at a confidence threshold of 0.8–1.

### Microbiome Difference Analysis and Functional Prediction

Optimized sequences were subjected to Operational Taxonomic Unit (OTU) clustering analysis and species taxonomic analysis. Krona (version 2.6)[21] was employed to visualize the abundance statistics of each species classification. Species abundance stacked plots were generated using the ggplot2 package in R language, while species abundance heatmaps were plotted with the pheatmap package in R language. α-diversity metrics (Chao1, Shannon, Simpson, and Pielou’s evenness) were subjected to differential analysis using the Vegan package in R language. The Wilcoxon rank-sum test was used to evaluate differences in α-diversity among groups. Principal Coordinates Analysis (PCoA) based on Bray-Curtis distance was performed using the Vegan package in R language, and the resulting plots were generated with the ggplot2 package. Linear Discriminant Analysis Effect Size (LEfSe) [22] (LDA > 3, p < 0.05) was applied to screen for microbial taxa with significant differences among groups, with the screening criteria set as LDA > 3 and p < 0.05. PICRUSt2 (version 2.5.3)[23] (version 2.5.3) was used to predict the KEGG (Kyoto Encyclopedia of Genes and Genomes) metabolic pathways. The raw sequence data reported in this paper have been deposited in the Genome Sequence Archive (Genomics, Proteomics & Bioinformatics 2025) in National Genomics Data Center (Nucleic Acids Res 2025), China National Center for Bioinformation / Beijing Institute of Genomics, Chinese Academy of Sciences (GSA: CRA043556) that are publicly accessible at https://ngdc.cncb.ac.cn/gsa.Users can access the data through the provided accession number following the GSA data access policies[24, 25].

## Results

### General Characteristics

A total of 13 subjects were enrolled in this study, consisting of 7 patients in the androgenetic alopecia (AGA) group and 6 subjects in the healthy control (HC) group. No significant differences were observed between the two groups in terms of age, sex distribution, body mass index (BMI), smoking rate, or alcohol consumption rate (all *P* > 0.05), ensuring the comparability of the two groups. Notably, the AGA group exhibited significantly more frequent hair washing, higher incidence of staying up late, stronger preference for greasy food, and higher rate of insufficient fruit and vegetable intake compared with the HC group (all P < 0.05). See Table 1.

**Table 1.** Comparison of general characteristics between androgenetic alopecia (AGA) and healthy control (HC) groups.

| Variable | AGA group (n=7) | HC group (n=6) | P-value |
| --- | --- | --- | --- |
| Age, years | 24.00 (21.00, 30.00) | 32.50 (29.00, 36.00) | 0.16 |
| Sex, male/female | 3/4 | 0/6 | 0.105 |
| BMI, kg/m<sup>2</sup> | 22.92±2.81 | 21.09±0.97 | 0.17 |
| Hair washing interval, days | 2.00±0.53 | 3.08±0.20 | <0.001* |
| Staying up late, n (%) | 6 (85.7) | 2 (33.3) | 0.047* |
| Smoking, n (%) | 2 (28.6) | 0 (0.0) | 0.487 |
| Alcohol drinking, n (%) | 1 (14.3) | 0 (0.0) | 1 |
| Dietary habits |  |  |  |
| Preference for sweet food | 2(28.6) | 1(16.7) | 1 |
| Preference for salty food | 1(14.3) | 0(0.0) | 1 |
| Preference for greasy food | 4(57.1) | 0(0.0) | 0.035* |
| Preference for spicy food | 5(71.4) | 3(50.0) | 0.592 |
| Insufficient fruit and vegetable intake | 4(57.1) | 0(0.0) | 0.035* |

Data are presented as median (interquartile range), mean ± standard deviation, or number (percentage). The Wilcoxon rank-sum test or Chi-square test was used for group comparisons. \* $P < 0.05$ indicates a significant difference.

Among the 7 AGA patients, the most common site of hair loss was the vertex (4 cases, accounting for 57.14%), followed by the hairline (3 cases, accounting for 42.86%). Regarding scalp symptoms, scalp redness was the most prevalent (5 cases, 71.43%), followed by scalp itching and excessive oiliness (4 cases each, 57.14% respectively), while increased dandruff was rarely observed (1 case, 14.29%). Additionally, a positive family history of AGA was found in 3 patients (42.86%). See Table 2.

**Table 2.** Scalp characteristics and family history in the AGA group.

| characteristic | No. of cases (n) | Proportion (%) |
| --- | --- | --- |
| Hair loss site |  |  |
| Hairline | 3 | 42.86 |
| Vertex | 4 | 57.14 |
| Scalp characteristics |  |  |
| Itching | 4 | 57.14 |
| Oiliness | 4 | 57.14 |
| Scalp redness | 5 | 71.43 |
| Increased dandruff | 1 | 14.29 |
| Family history | 3 | 42.86 |

### Bacterial Community Composition

At the genus level, the scalp bacterial community composition differed significantly between the AGA and HC groups (Fig 1a). The HC group showed a highly concentrated community structure, with *Staphylococcus* accounting for more than 50% of the total relative abundance. In contrast, the AGA group exhibited a marked reduction in *Staphylococcus* abundance, accompanied by an increase in opportunistic pathogenic genera such as *Acinetobacter* and *Pseudomonas*, resulting in a more dispersed and heterogeneous community structure.

**Fig 1.**
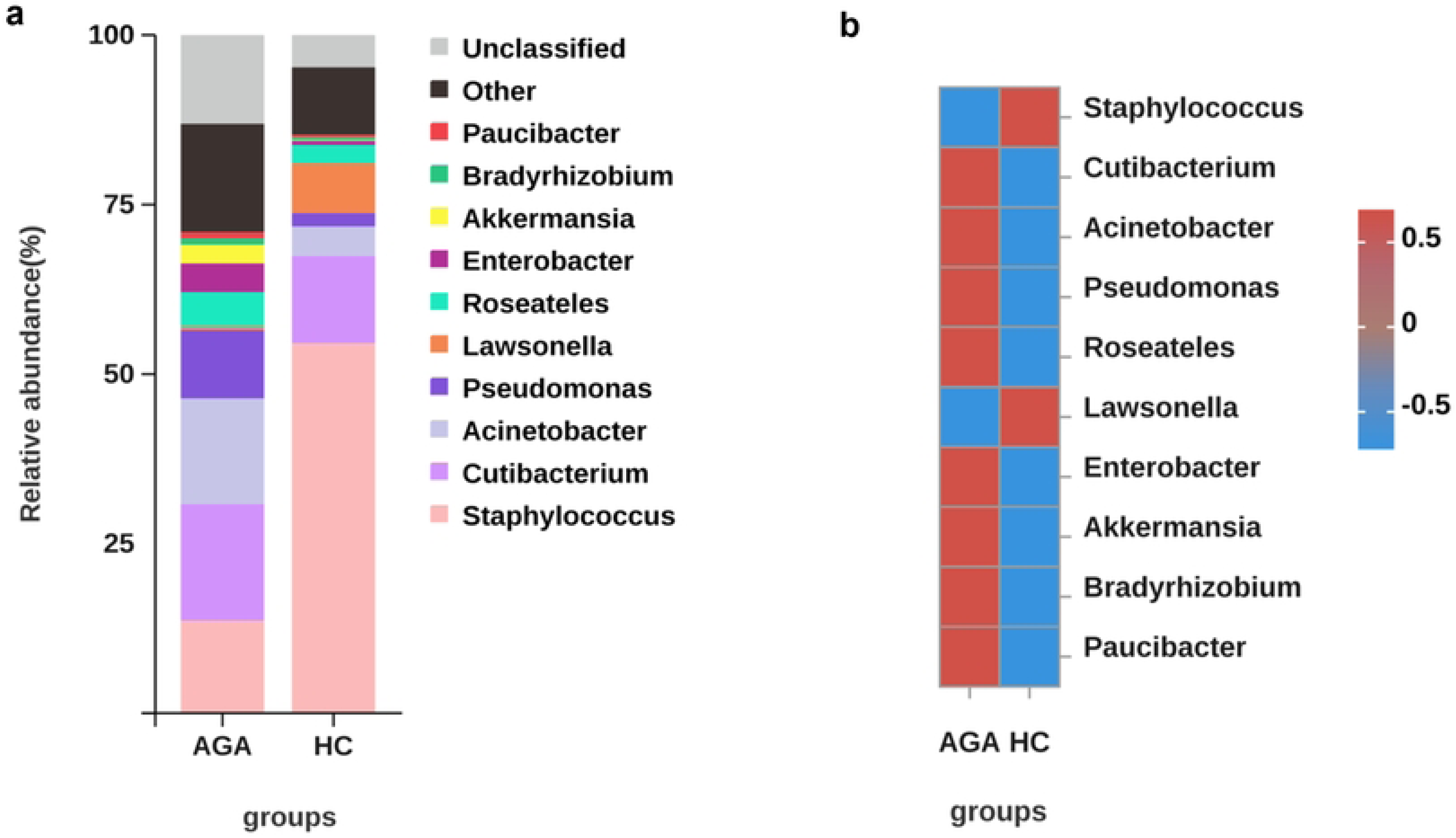
Genus-level scalp bacterial community composition and differential abundance between the AGA and HC groups. (a) Stacked bar plot showing the relative abundance of the top bacterial genera in the androgenetic alopecia (AGA) and healthy control (HC) groups. (b) Heatmap of normalized relative abundance of bacterial genera showing distinct enrichment patterns between groups. Red indicates higher relative abundance, while blue indicates lower relative abundance.

The normalized heatmap further confirmed these distinct enrichment patterns (Fig 1b). The HC group was characterized by the enrichment of *Staphylococcus* and *Lawsonella*. Meanwhile, the AGA group was significantly enriched with multiple genera, including *Cutibacterium* (formerly *Propionibacterium acnes*), *Acinetobacter*, *Pseudomonas*, *Roseateles*, *Enterobacter*, *Akkermansia*, *Bradyrhizobium*, and *Paucibacter*.

### Alpha-Diversity of Scalp Bacterial Microbiota

Alpha-diversity was assessed using observed features, Chao1, Shannon, Simpson, and Pielou evenness indices (Fig 2). The Chao1 index revealed no significant difference in species richness between the androgenetic alopecia (AGA) and healthy control (HC) groups (*p* = 0.9452). In contrast, the Shannon index (*p* = 0.0047), Simpson index (*p* = 0.0082), and Pielou(*p* = 0.0012) evenness index showed highly significant differences, indicating a significant increase in microbial diversity in the AGA group compared with the HC group.

**Fig 2.**
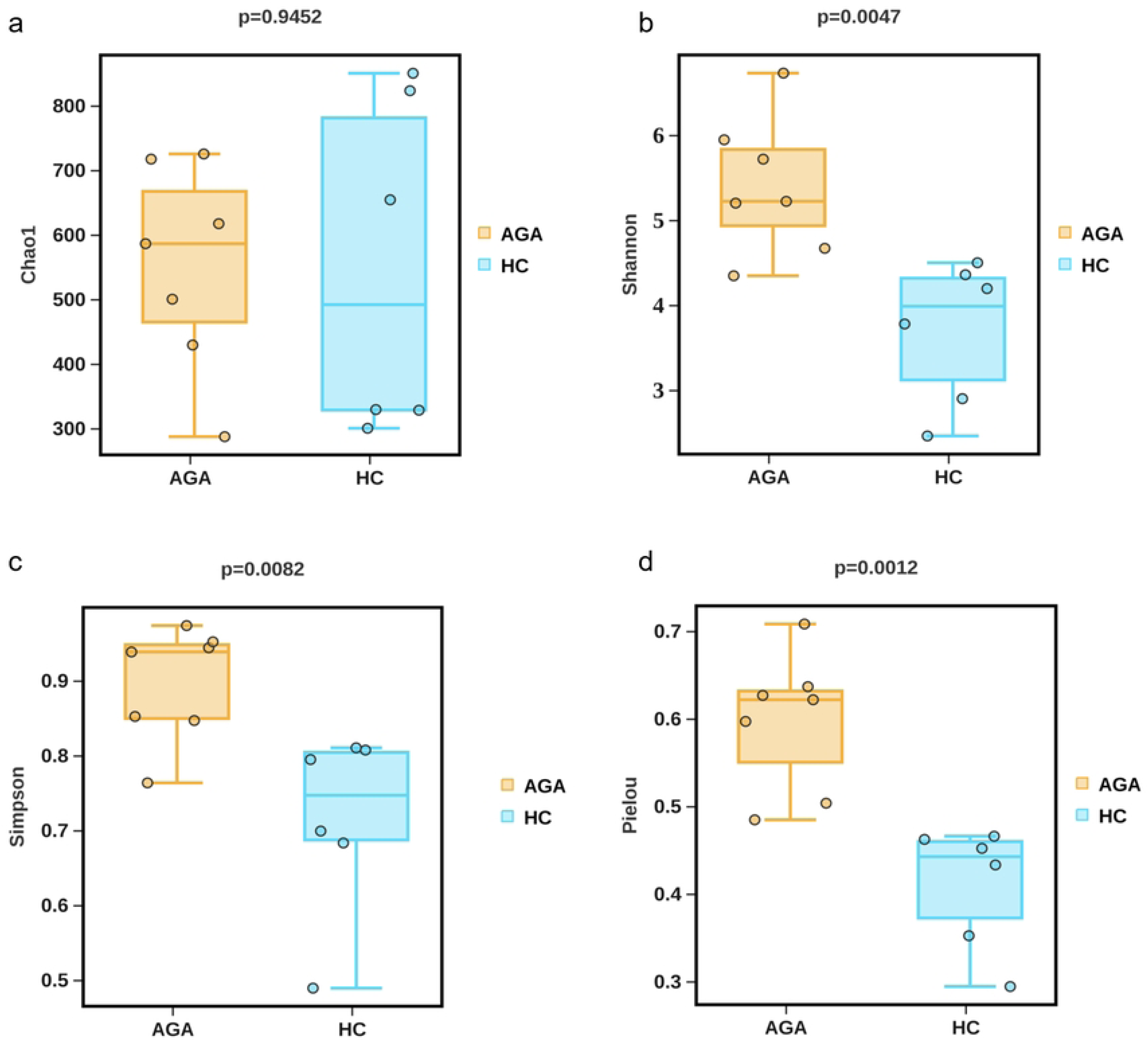
The alpha-diversity analysis of scalp bacterial microbiota in each group. These box plots show the alpha–diversity estimation scores of each group, which were calculated using (a) Chao1 index ; (b) Shannon’s index; (c) Simpson’s index; and (d) Pielou’s evenness.Group comparisons were evaluated using statistical analysis (Wilcoxon rank-sum test (Mann–Whitney U test)). Compared with healthy controls, the Chao index showed no significant difference in the androgenetic alopecia group, whereas the Shannon, Simpson and Pielou indices were significantly higher.

### Beta-Diversity of Scalp Bacterial Microbiota

The β-diversity PCoA analysis based on 16S rRNA gene sequencing revealed significant differences in microbial community structure between the androgenetic alopecia (AGA) and healthy control (HC) groups (Fig 3a). PCo1 and PCo2 explained 35.27% and 18.27% of the total variation, respectively. The two groups were clearly separated along the PCo1 axis without obvious overlap, indicating fundamental differences in microbial community composition patterns between healthy and alopecic states.

**Fig 3.**
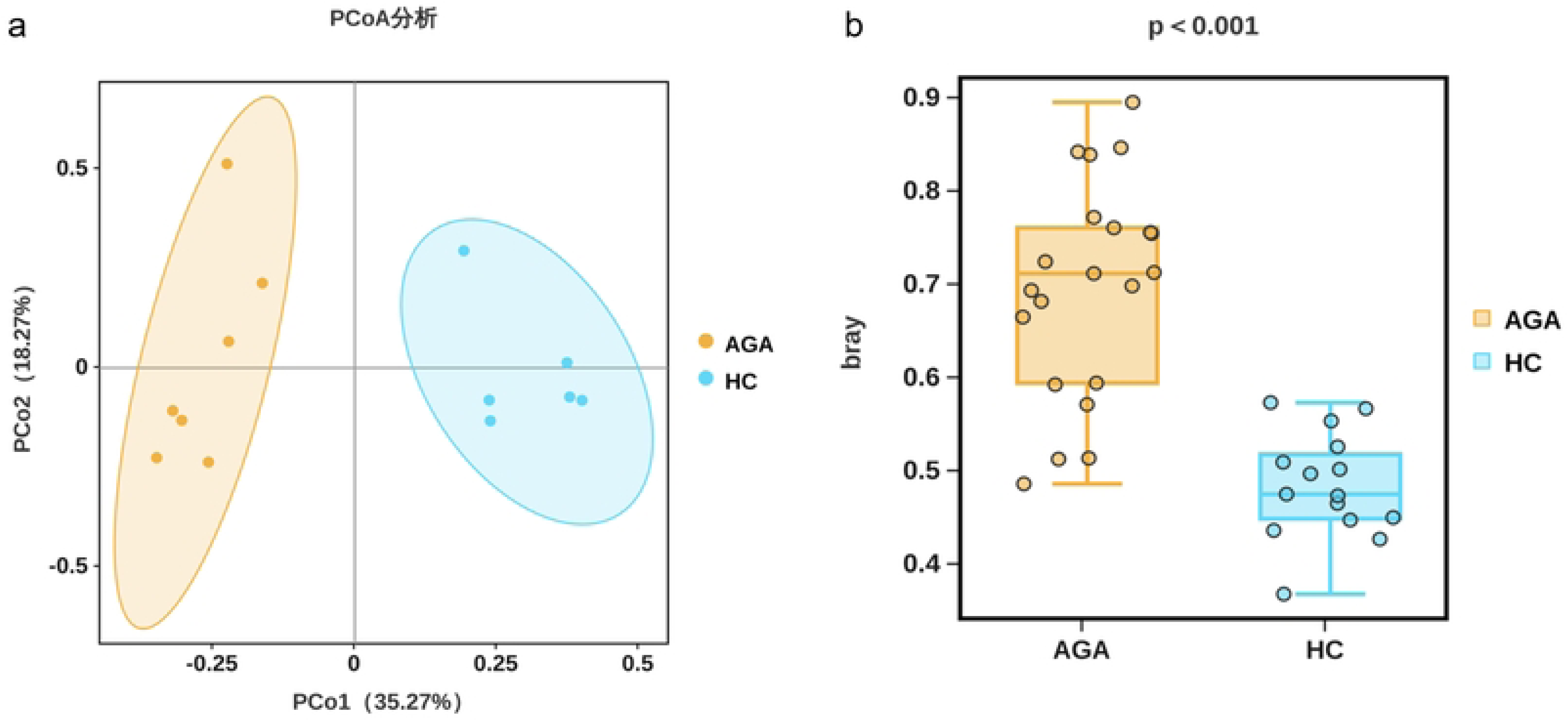
PCoA analysis and the Wilcoxon test revealed highly significant differences in microbial community structure between the AGA and HC groups. PCoA based on Bray-Curtis distance (a) showed obvious separation between the two groups. The Wilcoxon rank-sum test (b) confirmed a highly significant difference in Bray-Curtis distance between groups (*p* < 0.001)), suggesting substantial distinctions in scalp microbial composition between AGA patients and healthy individuals.

Furthermore, the Bray–Curtis distance confirmed that the intra-group distance was significantly higher in the AGA group than in the HC group (*p* < 0.001), suggesting a remarkable increase in community dispersion, enhanced inter-individual heterogeneity, and fundamental restructuring of the microbial community structure in AGA patients (Fig 3b).

### Genus- and Species-Level Differences in the Scalp Microbiota Between AGA Patients and Healthy Controls

Genus- and Species-Level Differences in the Scalp Microbiota Between AGA Patients and Healthy Controls Significant differences in microbial community composition were observed between the androgenetic alopecia (AGA) group and healthy control (HC) group at both the bacterial genus and species levels (Fig 4). At the genus level, the relative abundances of *Staphylococcus* and *Lawsonella* were significantly higher in the HC group than in the AGA group (*p* < 0.05; Fig 4a). At the species level, multiple *Acinetobacter* species, including *Acinetobacter radioresistens*, *Acinetobacter guillouiae*, *Acinetobacter sp. ACNIH1*, and *Acinetobacter pittii*, were significantly enriched in the AGA group compared with HC (*p* < 0.05; Fig 4b). Similarly, *Pseudomonas parafulva*, *Helicobacter typhlonius*, *Veillonella parvula*, and *Akkermansia muciniphila* were also significantly more abundant in AGA patients (*p* < 0.05; Fig 4b).

**Fig 4.**
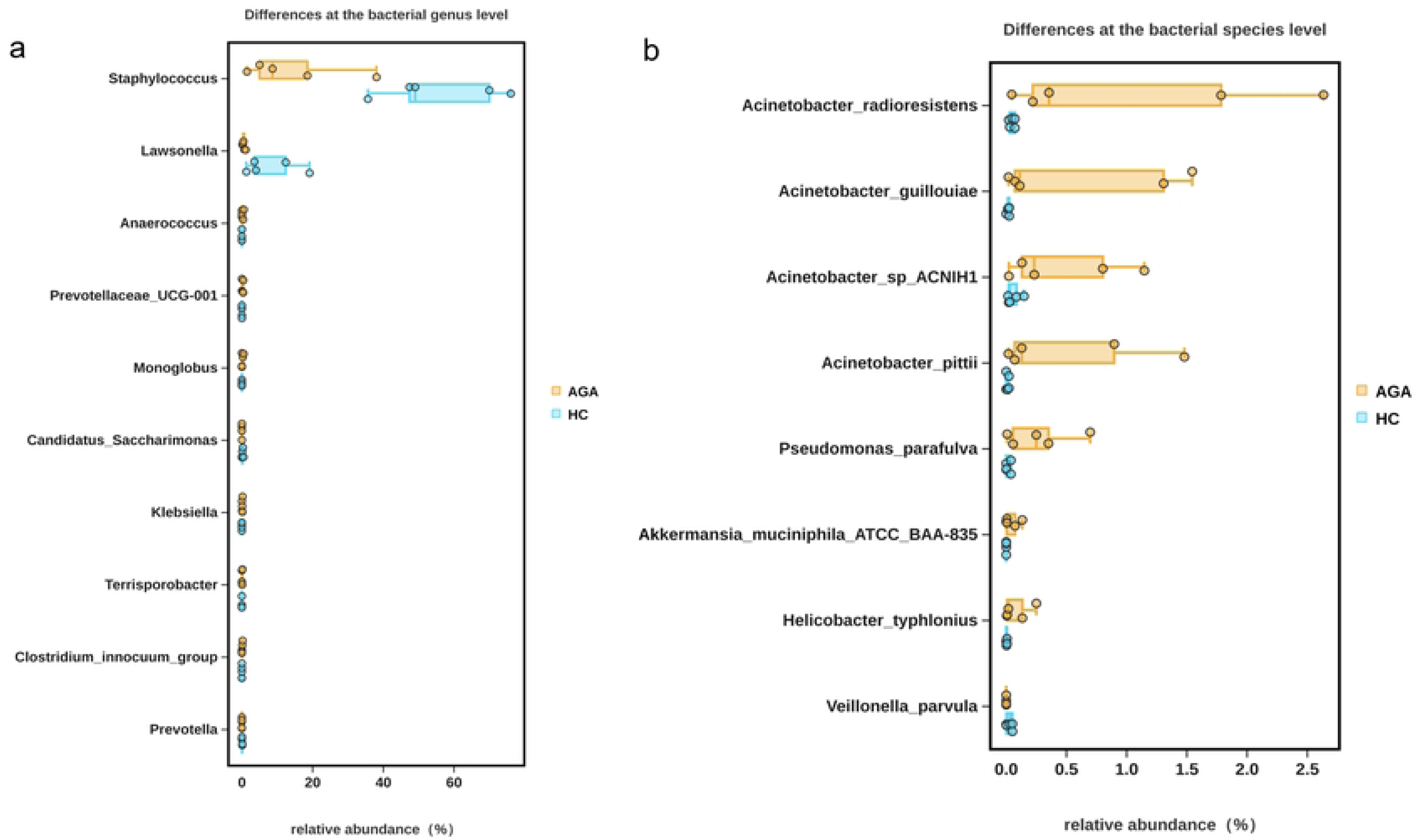
Relative abundance of significantly different bacterial taxa in AGA patients and healthy controls (HC). (a) Genus-level differences. (b) Species-level differences. Only taxa with statistically significant differences (p < 0.05, Wilcoxon rank-sum test) are shown. AGA, androgenetic alopecia; HC, healthy controls.

### Significant Microbiota Biomarkers Identified Using LEfSe Analysis

Linear discriminant analysis effect size (LEfSe) revealed significant differences in the scalp microbial signatures between AGA patients and healthy controls (LDA score > 3.5, *p* < 0.05; Fig 5). The AGA group was characterized by a distinct profile of inflammation-associated taxa, including *Pseudomonadota* (LDA = 5.24), *Gammaproteobacteria* (LDA = 5.23), *Acinetobacter* (LDA = 4.74), and *Pseudomonas* (LDA = 4.62), suggesting a marked inflammatory dysbiosis in the scalp microenvironment under AGA conditions. In contrast, the HC group was significantly enriched with health-related commensal bacteria, such as *Bacillota* (LDA = 5.26), *Staphylococcales* (LDA = 5.29), and *Staphylococcus* (LDA = 5.28). These findings demonstrate a clear “inflammatory-type” versus “health-type” dichotomy in the scalp microbiota structure between the two groups, providing a theoretical foundation for exploring the role of dysbiosis in AGA pathogenesis and potential microecological intervention strategies.

**Fig 5.**
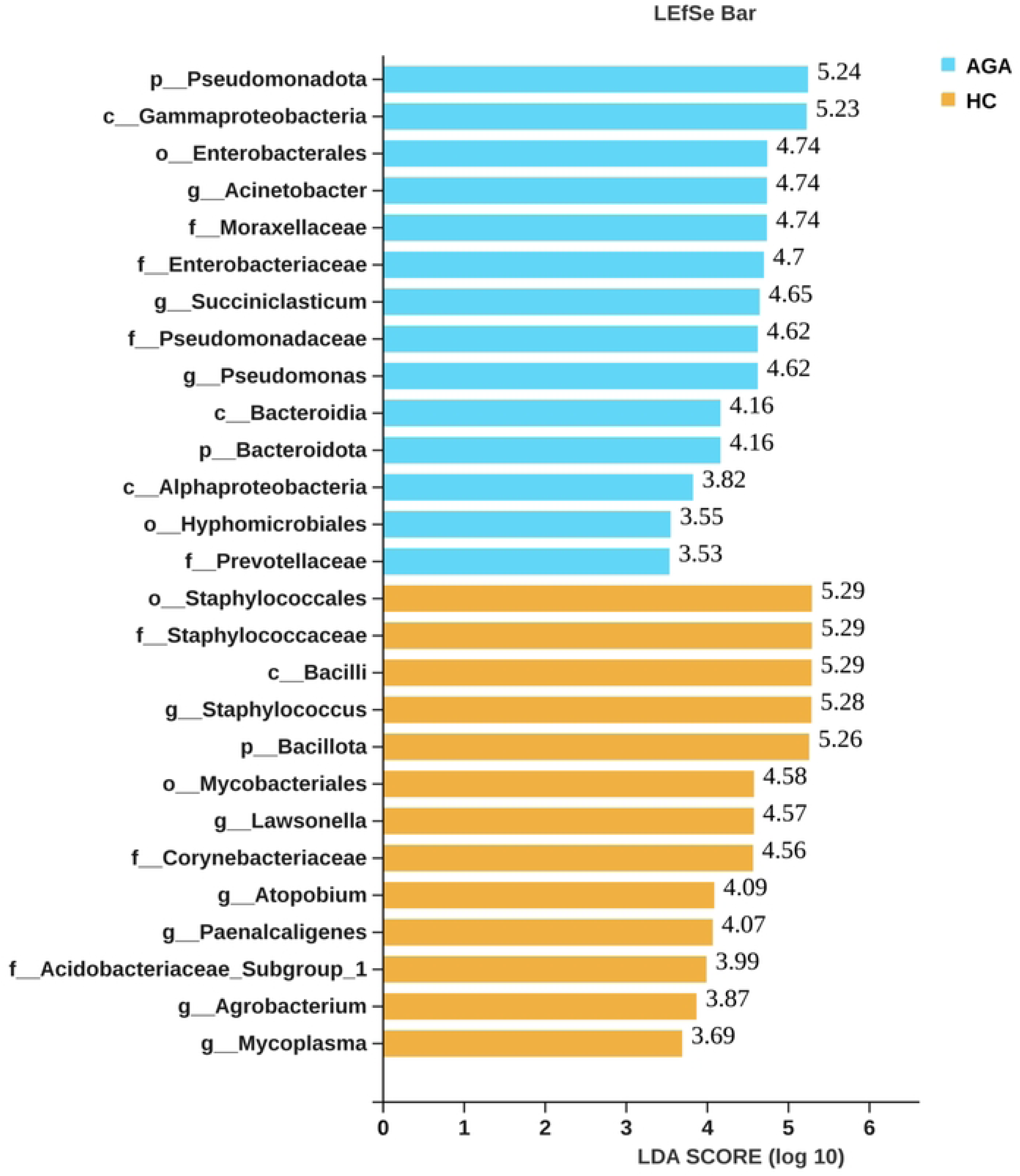
Linear discriminant analysis effect size (LEfSe) analysis of bacterial taxa differentiating the AGA and HC groups. Taxa with an LDA score > 3.0 (log10) and *p* < 0.05 are displayed. Blue bars represent taxa significantly enriched in the AGA group, while orange bars represent taxa significantly enriched in the HC group. AGA, androgenetic alopecia; HC, healthy controls.

### Functional Prediction of the Scalp Microbiota

KEGG level 1 functional pathway analysis revealed that, compared with the HC group, the AGA group exhibited significantly higher abundances of pathways related to carbohydrate metabolism, metabolism of cofactors and vitamins, amino acid metabolism, metabolism of terpenoids and polyketides, xenobiotics biodegradation and metabolism, lipid metabolism, folding, sorting and degradation, cell motility, signal transduction, and the immune system (*p* < 0.05). In contrast, only the infectious diseases pathway was significantly enriched in the HC group compared with AGA (*p* < 0.05; Fig 6).

**Fig 6.**
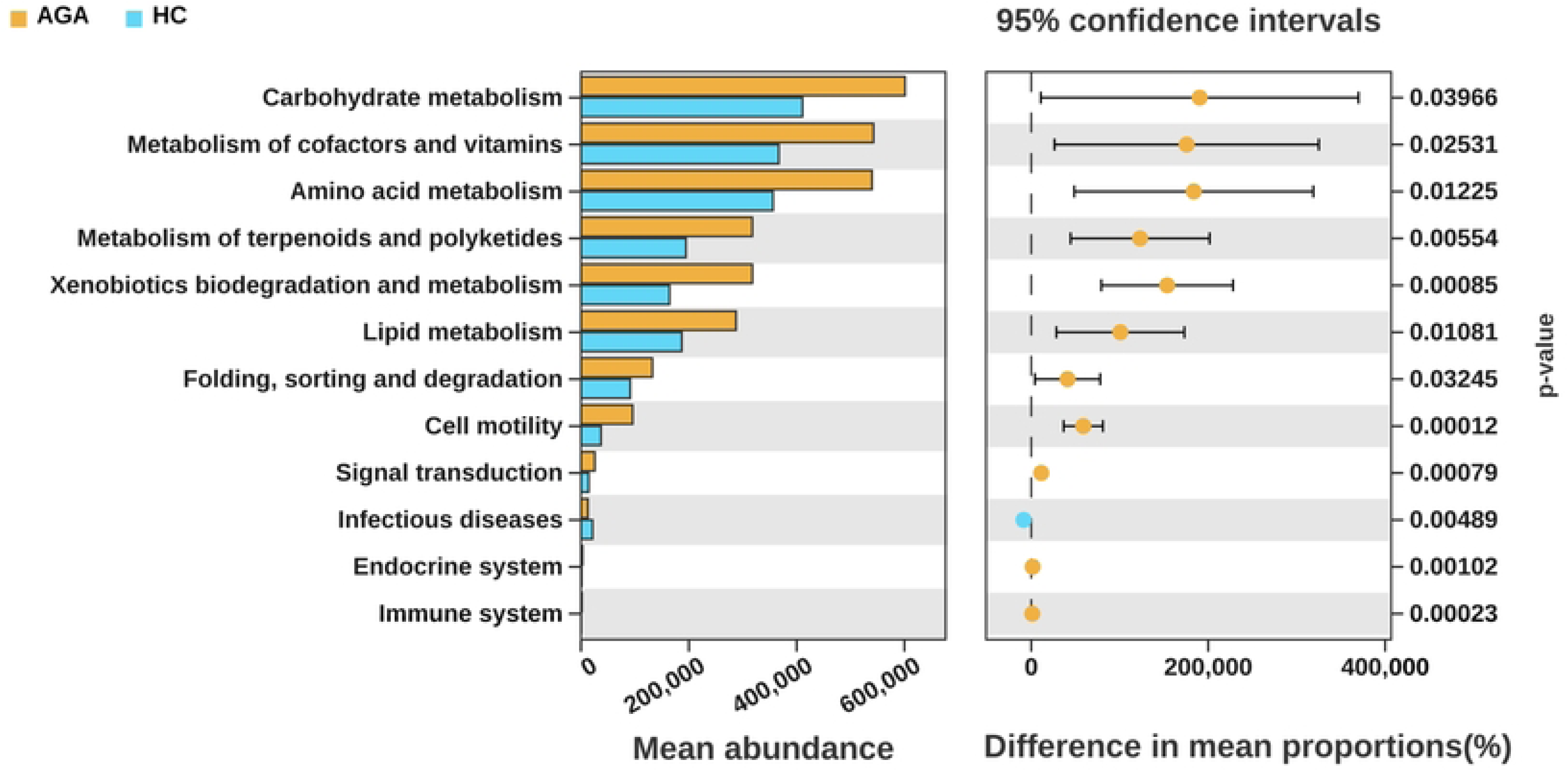
Functional prediction of scalp microbiota based on PICRUSt2 analysis, showing significantly different KEGG level 1 pathways between AGA patients and healthy controls (HC). Left panel: Mean abundance of predicted pathways in the AGA (orange) and HC (blue) groups. Right panel: Differences in mean proportions (%) with 95% confidence intervals and corresponding *p*-values. Only pathways with statistically significant differences (*p* < 0.05) are displayed. AGA, androgenetic alopecia; HC, healthy controls.

## Discussion

In this study, we employed 16S rRNA high-throughput sequencing to systematically investigate the structure, diversity patterns, and functional profiles of the scalp bacterial community in patients with androgenetic alopecia (AGA) and healthy controls (HC). Our results demonstrate that AGA patients exhibit a distinct scalp dysbiosis phenotype, characterized by reduced commensal bacteria, abnormally elevated α-diversity, diminished community stability, significant enrichment of pro-inflammatory taxa, and strong associations with unhealthy lifestyle factors. These findings provide novel insights into the microbial pathogenesis of AGA and offer experimental evidence supporting the scalp microbiota as both potential disease biomarkers and therapeutic targets.

Regarding α-diversity, the Chao index showed no significant difference between the two groups, suggesting comparable species richness. However, the Shannon index, Simpson index, and Pielou’s evenness index were all significantly elevated in the AGA group (all *P* < 0.01), in line with previous reports[12].These results indicate that the scalp microbial community in AGA is not characterized by simple species loss, but undergoes profound ecological restructuring. A healthy scalp is usually dominated by a limited set of core commensal bacteria, forming a compact and highly stable community structure. In pathological states, by contrast, scalp barrier impairment and shifts in the local microenvironment enable invasion and proliferation of low-abundance non-indigenous bacteria. Interactions among these non-commensal taxa further dilute the dominance of core commensals, increase community heterogeneity, and exacerbate disease development and progression[26].This pattern closely mirrors microbial dysbiosis observed in other skin and scalp disorders, including acne, atopic dermatitis, and alopecia areata, supporting the concept that abnormal diversity reflects ecological imbalance[27–31].

β-diversity analysis further verified significant structural separation between the AGA and HC groups. Importantly, the within-group Bray–Curtis distance was markedly larger in the AGA group (*P* < 0.001), indicating significantly increased inter-individual microbial heterogeneity and collapsed community stability in AGA patients. Such high dispersion implies that during alopecia progression, the scalp microbiota loses its homeostatic structure and shifts toward a highly unstable pathological state. Collectively, these diversity profiles define a consistent and reproducible dysbiosis signature of the scalp in AGA.

In terms of community composition, the healthy scalp was dominated by *Staphylococcus* as the predominant taxon (relative abundance > 50%)[32]; In contrast, the relative abundance of *Staphylococcus* was significantly decreased in the AGA group, whereas opportunistic pathogens such as *Cutibacterium* (formerly *Propionibacterium*), *Acinetobacter*, and *Pseudomonas* were markedly enriched[1]. LEfSe analysis identified *Bacillota* and *Staphylococcus* as signature taxa of the healthy scalp, while *Pseudomonadota*, *Gammaproteobacteria*, *Acinetobacter*, and *Pseudomonas* served as robust biomarkers for the AGA group. On the healthy scalp surface, *Staphylococcus* produces multiple antimicrobial peptides including bacteriocins, defensins, LL-37, and REG3A. Meanwhile, its lipoteichoic acid can activate the TLR2 signaling pathway in keratinocytes and enhance the expression of tight junction proteins, thereby playing a crucial role in maintaining the integrity of skin structure, regulating local immunity, and inhibiting the proliferation of pathogenic bacteria[33, 34]. The reduced abundance of *Staphylococcus* indicates impaired scalp defense, which creates a favorable environment for the colonization of opportunistic pathogens. *Cutibacterium* helps maintain the weakly acidic pH of the healthy scalp by releasing free fatty acids; however, its overgrowth leads to excessive accumulation of free fatty acids and subsequent damage to the skin barrier[35, 36]. Furthermore, *Cutibacterium* can synthesize multiple enzymes involved in porphyrin metabolism, triggering chronic perifollicular inflammation, accelerating hair follicle miniaturization, and promoting the progression of alopecia[1, 37]. Dysregulation of *Cutibacterium* has also been associated with acne vulgaris, dandruff, and seborrheic dermatitis [38–40].

KEGG functional prediction demonstrated that numerous metabolic and functional pathways were significantly elevated in the AGA group. These enriched metabolic and immune-related pathways indicate that the scalp microenvironment in AGA patients exhibits an overall hypermetabolic and pro-inflammatory state. Upregulation of metabolic pathways reflects increased consumption of sebum-derived substrates by the scalp microbiota in AGA individuals, which may disrupt normal hair follicle growth and development [41–43], In line with these metabolic shifts, Kondrakhina et al. reported that serum levels of trace elements and vitamins were significantly lower in male AGA patients compared with healthy controls [44]. Enhanced cell motility and signal transduction further suggest increased microbial invasiveness within the scalp niche. In contrast, the infectious diseases pathway was more highly represented in the healthy control group, indicative of a more balanced and robust immune defense in the healthy scalp. Collectively, these functional alterations promote the generation of pro-inflammatory metabolites, compromise scalp barrier integrity, aggravate hair follicle injury, and ultimately accelerate the pathological progression of AGA [45].

The present study also revealed a close association between unhealthy lifestyles and scalp microbial dysbiosis.Compared with healthy controls, the AGA group exhibited significantly higher frequencies of hair washing, more frequent staying up late, a higher proportion of high-fat diets, and more insufficient fruit and vegetable intake (all *P* < 0.05)[46, 47]. Salem et al.[48] reported a higher prevalence of AGA in smokers than in non-smokers; however, this association was not observed in the current study, which may be attributed to the limited sample size.Lifestyle factors can directly or indirectly reshape microbial community structure by modulating sebum secretion, scalp oxidative status, local immunity, and the metabolic microenvironment of hair follicles[4, 49]. Our findings suggest that adverse lifestyles may contribute to AGA pathogenesis by exacerbating microbial dysbiosis, forming a sequential pathological cascade: unhealthy lifestyle → microbial dysbiosis → follicular inflammation → hair loss.These results provide a microbiological explanation for the epidemiological link between lifestyle and AGA, and support that lifestyle modification could serve as an adjuvant strategy to restore microbial balance in the early stages of AGA.

Several limitations in the present study should be acknowledged. First, given its cross-sectional observational design, causal inferences between microbial dysbiosis and AGA cannot be established; accordingly, it remains unclear whether the observed ecological imbalance represents a primary cause or a secondary consequence of hair follicle degeneration. Second, the relatively modest sample size may restrict the generalizability of our findings to broader populations. Third, the absence of longitudinal follow-up and targeted microbiota intervention trials precludes an evaluation of whether restoring microbial homeostasis can attenuate disease progression. Future investigations employing larger cohorts, prospective study designs, integrated multi-omics approaches, and topical microbial interventions are warranted to further elucidate causal relationships and advance clinical translation.

## Conclusions

In conclusion, the present study confirms that patients with androgenetic alopecia (AGA) exhibit distinct and distinguishable scalp microbial dysbiosis, which is characterized by a reduction in commensal bacteria (e.g., *Staphylococcus*), expansion of pro-inflammatory taxa (e.g., *Acinetobacter*, *Pseudomonas*, and *Cutibacterium*), abnormally elevated α-diversity (with significantly higher Shannon index, Simpson index, and Pielou’s evenness index), and increased community dispersion. Additionally, a significant association was observed between AGA and unhealthy lifestyle factors, including frequent hair washing, staying up late, high-fat diet, and insufficient intake of fruits and vegetables. Functional prediction based on KEGG pathways demonstrated that the dysbiotic microbiota in AGA patients exhibited enhanced metabolic and immune-related functional activities, which may be involved in the processes of perifollicular inflammation and hair follicle miniaturization. These findings refine the microbial pathogenesis model of AGA, supporting the potential of scalp microbiota as a biomarker for AGA risk assessment, early warning, and targeted intervention. Collectively, this study provides experimental evidence for the development of microecology-based prevention and treatment strategies for AGA, while also opening up a new direction for the precise and personalized management of hair disorders. It should be noted that due to the cross-sectional observational design and relatively limited sample size of this study, causal relationships between microbial dysbiosis and AGA cannot be definitively established, and the generalizability of the findings may be constrained. Future prospective cohort studies with larger sample sizes, integrated multi-omics approaches, and targeted microbial intervention trials are therefore warranted to further clarify the causal mechanisms and promote the clinical translation of these findings.

## Author Contributions

**Conceptualization:** Juntong Yu, Jiafeng Yang, Gang Deng.

**Methodology:** Yiwen He, Lulu Zhu.

**Software:** Yiwen He, Lulu Zhu.

**Validation:** Dingfeng Lv, Jiafeng Yang, Gang Deng.

**Formal Analysis**: Yiwen He, Lulu Zhu.

**Investigation:** Yiwen He, Lulu Zhu, Jinyu Wu, Jiale Jin.

**Resources:** Juntong Yu, Gang Deng.

**Data Curation:** Yiwen He, Lulu Zhu, Jinyu Wu, Jiale Jin.

**Writing—Original Draft:** Yiwen He.

**Writing—Review and Editing:** Dingfeng Lv, Gang Deng.

**Supervision:** Dingfeng Lv, Jiafeng Yang, Gang Deng.

**Funding Acquisition:** Juntong Yu, Gang Deng.

**Project Administration:** Juntong Yu, Gang Deng.

## Data Availability Statement

The raw sequence data have been deposited in the Genome Sequence Archive (GSA) under accession number CRA043556.

